# Types and timing of trauma exposure across the life course and maternal hypertension

**DOI:** 10.1101/2023.12.13.23299937

**Authors:** Kaitlyn K Stanhope, Vasiliki Michopoulos, Abigail Powers, Sheree L Boulet, Michael R Kramer, Shakira F Suglia

## Abstract

**Objective:** To estimate associations between types and timing (first occurrence and time since) of trauma exposure and hypertension experienced during pregnancy in a safety-net hospital in Atlanta, Georgia.

**Methods:** Participants completed a 14-item trauma screener. We linked that information to data from the medical record on hypertension (inclusive of chronic hypertension, gestational hypertension, or preeclampsia). We fit logistic regression models and used the estimates to calculate risk ratios for each trauma type and each critical window (0-9 years, 10-19, and 20+). We fit unadjusted models and adjusted for age, parity, and education.

**Results:** We included 704 individuals with a delivery within 12 months of screening. The majority (94%, 661) reported at least one traumatic event, most commonly witnessing violence (79.4%). Overall, 18% experienced gestational hypertension, 10.8% chronic hypertension, and 11.9% preeclampsia. Among individuals who reported trauma, 31.5% screened positive for probable posttraumatic stress disorder and 30.9% for probable depression compared to 0 and 2.3% among those without reported trauma. No trauma type (violence, witnessing violence, non-interpersonal, or sexual assault) was associated with increased hypertensive risk, regardless of timing. Similarly, time between trauma and delivery (0-3 years, 3-10 years, 10+ years) was not associated with increased hypertensive risk.

**Conclusions:** In this sample with a high trauma and hypertension burden, trauma was not associated with elevated risk of hypertension during pregnancy, despite a high burden of PTSD and depressive symptoms among people with trauma exposure. Future research should consider how community-level exposure may modify the impact of trauma on adverse pregnancy outcomes.

---

Exposure to trauma across the life course is linked to poor cardiovascular health including atherosclerotic cardiovascular disease, hypertension, and diabetes.^1^ Trauma exposure results in acute and long term physiologic changes, including shifts in vascular and neuroendocrine function.^2–4^ People living in poverty and of minoritized racial, ethnic, and gender identities experience disproportionately high rates of stress and trauma compared to white and high income people.^5–9^ While considerable evidence supports a relationship between trauma and cardiovascular disease in mid and late life, the relationship between trauma and poor cardiovascular health during other life stages, including pregnancy, remains unclear.

Increasingly, public health practitioners and clinicians are interested in the relationship between trauma exposure and cardiovascular risk during pregnancy. Pregnancy complications have been framed as a “window” to future cardiovascular disease.^10,11^ The physiologic response to pregnancy, including whether or not the pregnant person develops a hypertensive disorder of pregnancy or gestational diabetes, is thought to mirror physiologic responses to aging.^12^ Supporting this framework is the consistent evidence that people who experience hypertensive disorders of pregnancy are at increased risk of hypertension and experience an earlier onset of hypertension, compared to parous counterparts with uncomplicated pregnancies.^10,13^ Trauma exposure may result in poor cardiovascular health through changing behaviors, physiological shifts, epigenetic ageing, or the development of psychiatric symptoms.^1,14,15^ Understanding if and how trauma is related to hypertension during pregnancy may inform secondary prevention efforts for cardiovascular disease in people who have experienced trauma and our understanding of the mechanisms through which trauma increases cardiovascular risk.

To date, studies of trauma and hypertension during pregnancy have focused on trauma experienced during childhood, with mixed results. In a single-center study in the United States, Jasthi et al. (2023) identified a two-fold excess risk of hypertensive disorders of pregnancy among individuals who reported four or more adverse childhood experiences (ACEs) and screened positive for depression or anxiety during pregnancy compared to those with mental health symptoms but no ACEs.^16^ In a study among the Hispanic Community Health Study/Study of Latinos, the authors did not observe any association between accumulation of ACEs, and HDP or GDM.^17^ Neither of these studies accounted for trauma exposure occurring during adulthood, which may mediate part of the association between childhood trauma and maternal risk.

The Weathering Hypothesis posits that the accumulation of stress exposure results in elevated risk of poor pregnancy outcomes.^18,19^ Many authors have operationalized this as a count of events, regardless of when they happened in time. Friedman et al. compared model fit using a timing-based model and count-based definition of trauma and found that the type and number of traumatic events experienced in childhood was the strongest predictor of women’s heart disease risk.^20^ Alternately, individuals who experience trauma during adulthood, more proximate to conception, may experience acute psychiatric and physical consequences, possibly impacting pregnancy outcomes.^9^

In the current study, we focus on the potential association between characteristics of trauma exposure (type and timing) and hypertensive disorders of pregnancy in people seeking care at a safety-net hospital in Atlanta, Georgia. We hypothesize that more severe trauma exposure (e.g., directly experiencing violence or sexual assault) experienced in childhood will have the strongest association with hypertensive disorders of pregnancy, followed by the most recent trauma exposure.

## Methods

### Study Setting and Population

We used data from The Grady Trauma Project (GTP), conducted in Grady Memorial Hospital in Atlanta, Georgia. The GTP includes multiple substudies, two of which are included in this sample. The first is a long-running cross-sectional survey which recruited individuals in the waiting room of primary care, diabetes, and the Women’s clinic from 2012-2020.The second recruited individuals during pregnancy and followed them through three months postpartum from 2017-2022. For both, we linked information from the survey to the electronic medical record creating a cohort. For the cross-sectional GTP sample, we included deliveries to participants within twelve months following their survey. For the prospective GTP sample, we identified the index delivery linked to the pregnancy for which individuals were recruited (Supplemental Figure 1). Notably, the birthing population served by Grady faces a high burden of both medical comorbidities and ongoing social risks during their pregnancy compared to national averages.^21,22^ This study was approved by the Emory Institutional Review board (STUDY00001471). All participants consented to participation in the Grady Trauma Project.

### Measures

Participants reported trauma on the “Traumatic Events Inventory”, a 14-item scale which asks about the occurrence of traumatic events and the first age at which they occurred. The TEI includes the following domains: experiencing natural disaster, a serious accident or injury, a sudden, life-threatening illness, military combat, witnessed the murder of a close friend/family, experiencing attack with or without a weapon by an intimate partner or other person, witnessing a friend/family member being attacked with or without a weapon, witnessing violence between caregivers (in childhood), physical maltreatment in childhood, emotional maltreatment in childhood, sexual abuse in childhood, adolescence, or adulthood, and other trauma (open-ended).^23,24^ We classified traumatic events into four types: directly experienced violence, witnessed violence, sexual assault/abuse, and non-interpersonal trauma exposure. We also grouped events based on the age at which they first occurred: ages 0-9, 10-19, and 20+. For each event, we calculated a measure of “recency” or years between trauma and delivery. Participants who reported one or more traumatic events completed screening tools on post-traumatic stress disorder (PTSD) either modified PTSD Symptom Scale (mPSS)^25^ or Posttraumatic Stress Disorder Checklist for Diagnostic and Statistical Manual of Mental Disorders -5 (PCL-5).^25,26^ We used cut-offs to create binary measure of probable PTSD using guidance from the DSM IV-TR and 5, indicating symptoms from each cluster (three for DSM-IV and five for DSV-V).^25^

We used ICD-9 and ICD-10 codes as coded at the delivery hospitalization to identify hypertensive disorders including chronic hypertension, gestational hypertension, preeclampsia, or eclampsia at delivery. Evidence suggests that ICD codes accurately identify hypertensive disorders with positive predictive values between 85-90%.^27,28^ For the primary analysis, we collapsed all diagnoses into a single binary indicator of any maternal hypertension. We included the following covariates: maternal age at delivery, maternal education, and parity. Participants self-reported education in the survey.

### Analysis

We compared maternal characteristics by report of any traumatic event. We then fit multivariable logistic regression models to estimate the association between any traumatic event and trauma events in each category (directly experienced violence, witnessed violence, sexual assault/abuse, and non-interpersonal) separately. For models with trauma experienced at age 20+ as the exposure, we excluded birthing people less than 20. We used the “probratio” function in the R package “epitools” to transform beta estimates to risk ratios using maximum likelihood estimation.^29^ We conducted a complete case analysis, restricting to observations with complete data on the exposure, outcome, and included covariates. We fit unadjusted models and models adjusting for age at delivery (<20, 20-34, 35+), parity (primiparous or multiparous), and education (less than high school or high school graduate). We consider education as an incomplete measure of socioeconomic status. We also fit models estimating the association between trauma recency (time between first exposure to that type of trauma and delivery age) in each category.

We conducted two main sensitivity analyses. First, to account for differences in individuals who participated in the Grady Trauma Project and BUMPP studies and the overall Grady population, we fit inverse probability of participation rates.^30^ We considered age, parity, insurance, race/ethnicity, gravidity, chronic diabetes, and history of a mental illness diagnosis (ICD 9/10 code for alcohol or drug use disorder, mood disorder, PTSD, or anxiety) as important factors to use. We modeled the probability of participation using logistic regression and created stabilized weights. We repeated the primary models incorporating these weights. Second, we explored whether the overall observed associations differed by specific hypertensive diagnosis.

## Results

We included 704 people in the current sample (Table 1; Supplemental Figure 1). The majority of observations were dropped due to non-linkage with a delivery hospitalization (443 excluded), a delivery prior to completing the GTP survey (35 excluded), or a delivery more than 12 months following GTP (55 excluded). We further excluded 58 individuals missing the traumatic events inventory. Of those, the majority (94%, 661) reported at least one lifetime traumatic event. The majority of the study population graduated from high school (78.3%), reported no romantic partner (83.1%), and had Medicaid-insurance at delivery (94.2%). Just under a third of participants screened positive for probable PTSD (29.5%) and a similar proportion for depression (29.1%). Hypertensive disorders were common; over 40% of individuals had a diagnosis of one or more hypertensive disorders at delivery, most commonly gestational hypertension (18%) followed by chronic hypertension (10.8%).

**Figure 1.**
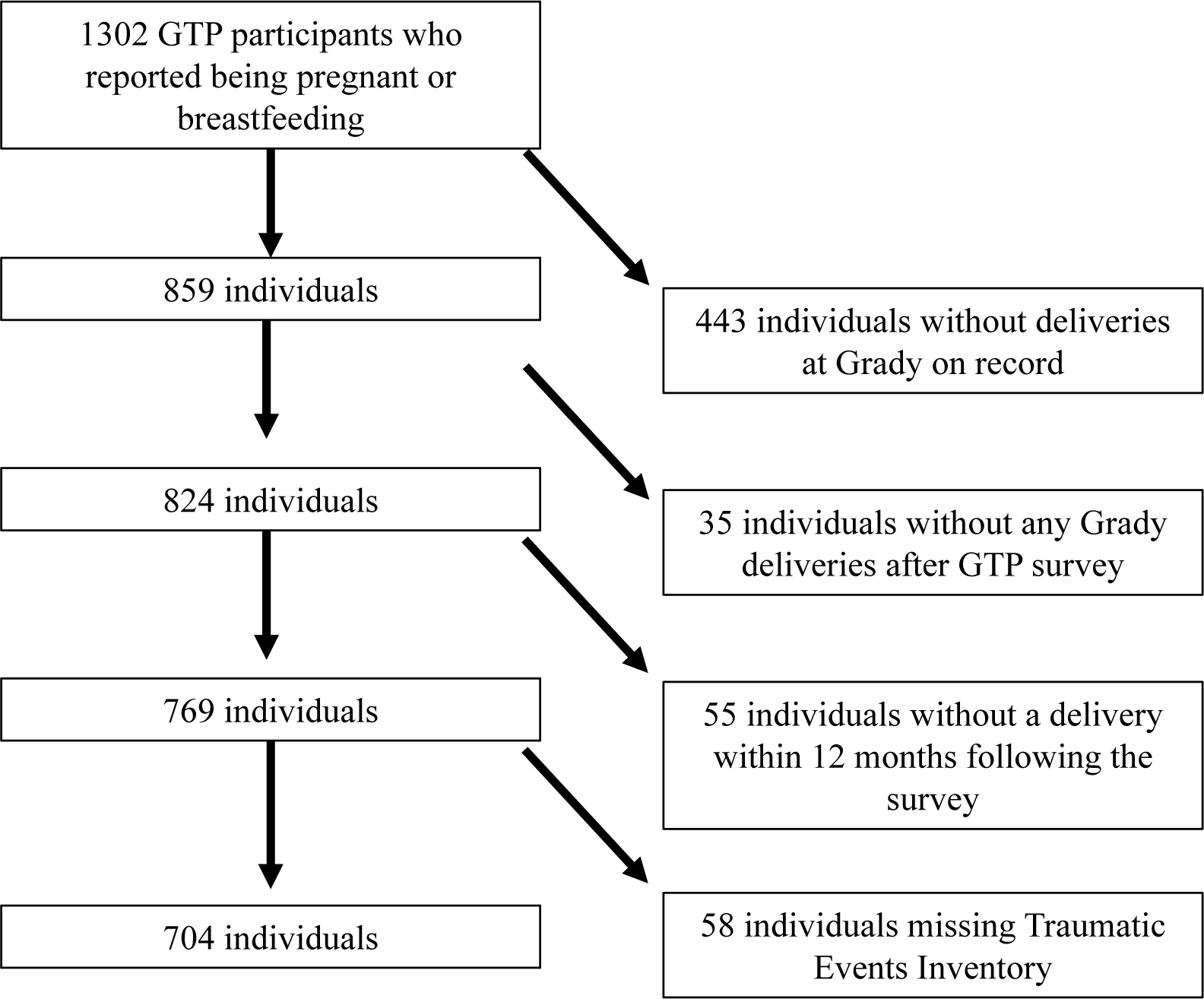
Study flow diagram showing exclusion at different stages, 704 included individuals with a delivery at Grady Memorial Hospital, 2011-2023, who participated in the Grady Trauma Project.

**Table 1.** Participant characteristics, stratified by lifetime trauma exposure, Grady Trauma Project, n = 704 individuals with a delivery between 2011-2023.

|  | One or more<br>traumatic events<br>(N=661) | No trauma reported<br>(N=43) | Overall<br>(N=704) |
| --- | --- | --- | --- |
| **Educational attainment** |  |  |  |
| Less than high school | 147 (22.2%) | 5 (11.6%) | 152 (21.6%) |
| High school graduate | 513 (77.6%) | 38 (88.4%) | 551 (78.3%) |
| Missing | 1 (0.2%) | 0 (0%) | 1 (0.1%) |
| **Age at delivery** |  |  |  |
| Mean (SD) | 26.8 (5.76) | 24.4 (5.31) | 26.7 (5.76) |
| Median [Min, Max] | 26.0 [16.0, 45.0] | 22.0 [18.0, 40.0] | 26.0 [16.0, 45.0] |
| **Monthly Income** |  |  |  |
| \$0-249 | 84 (12.7%) | 3 (7.0%) | 87 (12.4%) |
| \$250-499 | 67 (10.1%) | 9 (20.9%) | 76 (10.8%) |
| \$500-999 | 116 (17.5%) | 7 (16.3%) | 123 (17.5%) |
| \$1000-1999 | 200 (30.3%) | 6 (14.0%) | 206 (29.3%) |
| \$2000+ | 130 (19.7%) | 9 (20.9%) | 139 (19.7%) |
| Missing | 64 (9.7%) | 9 (20.9%) | 73 (10.4%) |
| **Reports romantic partner** |  |  |  |
| No | 553 (83.7%) | 32 (74.4%) | 585 (83.1%) |
| Yes | 108 (16.3%) | 11 (25.6%) | 119 (16.9%) |
| **Race/Ethnicity** |  |  |  |
| Non-Hispanic Black | 642 (97.1%) | 42 (97.7%) | 684 (97.2%) |
| Other | 15 (2.3%) | 1 (2.3%) | 16 (2.3%) |
| Missing | 4 (0.6%) | 0 (0%) | 4 (0.6%) |
| **Insurance during pregnancy** |  |  |  |
| Medicaid | 621 (93.9%) | 42 (97.7%) | 663 (94.2%) |
| Commercial insurance or uninsured | 40 (6.1%) | 1 (2.3%) | 41 (5.8%) |
| **Hypertension diagnosis at delivery** |  |  |  |
| Chronic hypertension | 71 (10.7%) | 5 (11.6%) | 76 (10.8%) |
| Superimposed Preeclampsia | 39 (5.9%) | 3 (7.0%) | 42 (6.0%) |
| Gestational hypertension | 117 (17.7%) | 10 (23.3%) | 127 (18.0%) |
| Preeclampsia without severe features | 15 (2.3%) | 0 (0%) | 15 (2.1%) |
| Preeclampsia with severe features or eclampsia | 27 (4.1%) | 0 (0%) | 27 (3.8%) |
| Normotensive | 392 (59.3%) | 25 (58.1%) | 417 (59.2%) |
| **Diabetes** |  |  |  |
| No diabetes | 603 (91.2%) | 37 (86.0%) | 640 (90.9%) |
| Chronic diabetes | 30 (4.5%) | 3 (7.0%) | 33 (4.7%) |
| Gestational diabetes | 28 (4.2%) | 3 (7.0%) | 31 (4.4%) |
| **Preterm birth, <37 weeks** |  |  |  |
| No | 551 (83.4%) | 36 (83.7%) | 587 (83.4%) |
| Yes | 106 (16.0%) | 7 (16.3%) | 113 (16.1%) |
| Missing | 4 (0.6%) | 0 (0%) | 4 (0.6%) |
| **Probable PTSD diagnosis** |  |  |  |
| No | 400 (60.5%) | 25 (58.1%) | 425 (60.4%) |
| Yes | 208 (31.5%) | 0 (0%) | 208 (29.5%) |
| Missing | 53 (8.0%) | 18 (41.9%) | 71 (10.1%) |
| **Probable depression diagnosis** |  |  |  |
| No | 409 (61.9%) | 41 (95.3%) | 450 (63.9%) |
| Yes | 204 (30.9%) | 1 (2.3%) | 205 (29.1%) |
| Missing | 48 (7.3%) | 1 (2.3%) | 49 (7.0%) |
| **Gestational Age at Entry to Prenatal Care** |  |  |  |
| Mean (SD) | 15.3 (7.49) | 13.5 (7.25) | 15.1 (7.48) |
| Median [Min, Max] | 13.0 [4.00, 38.0] | 11.0 [5.00, 31.0] | 13.0 [4.00, 38.0] |
| Missing | 8 (1.2%) | 0 (0%) | 8 (1.1%) |
| **Parity** |  |  |  |
| 0 prior births | 124 (18.8%) | 14 (32.6%) | 138 (19.6%) |
| 1 prior birth | 153 (23.1%) | 10 (23.3%) | 163 (23.2%) |
| 2+ prior births | 381 (57.6%) | 19 (44.2%) | 400 (56.8%) |
| Missing | 3 (0.5%) | 0 (0%) | 3 (0.4%) |
| **Childhood Trauma Questionnaire Score** |  |  |  |
| Mean (SD) | 42.0 (18.6) | 28.2 (5.58) | 41.2 (18.4) |
| Median [Min, Max] | 35.0 [25.0, 119] | 26.0 [25.0, 49.0] | 34.0 [25.0, 119] |
| Missing | 25 (3.8%) | 2 (4.7%) | 27 (3.8%) |
| **Number of prenatal care visits** |  |  |  |
| Mean (SD) | 8.07 (3.75) | 8.23 (3.65) | 8.08 (3.74) |
| Median [Min, Max] | 8.00 [0, 32.0] | 9.00 [2.00, 17.0] | 8.00 [0, 32.0] |

The most common type of trauma experienced was witnessing violence (79.4%), followed by non-interpersonal (66.6%), experiencing violence (60.2%), and sexual assault/abuse (38.9%) (Table 2). Participants reported experiencing trauma at young ages. On average, the mean age at first experience of sexual assault/abuse was 10.6 (SD: 5.3), witnessing violence 11.6 (SD: 6.2), experiencing violence 13.2 (SD: 6.7), and non-interpersonal trauma 14.3 (SD: 7.8).

**Table 2.**
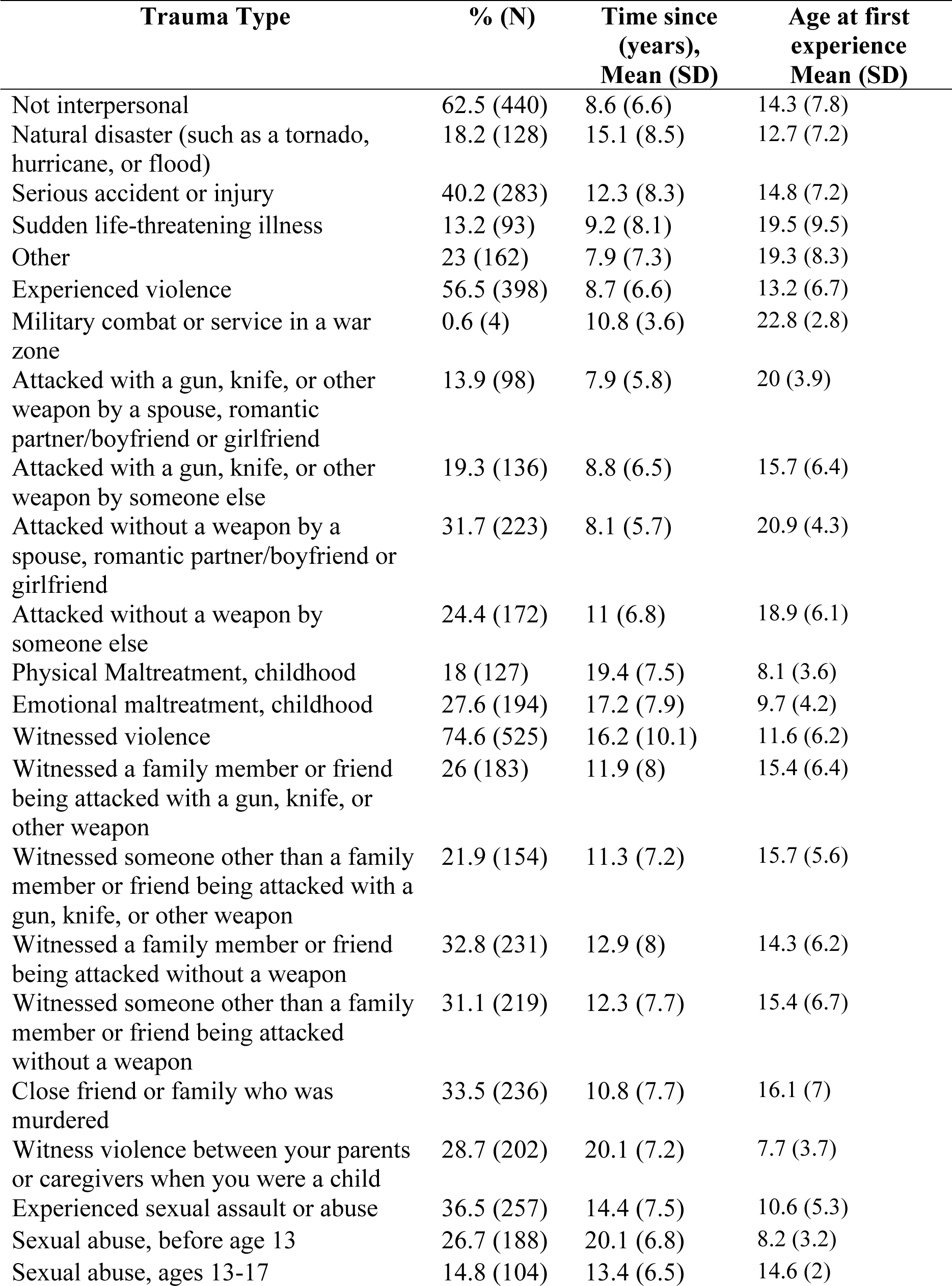
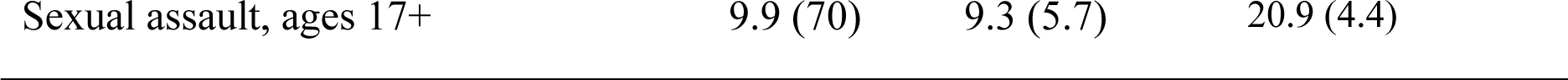
Reported traumatic events experienced or witnessed and mean years since, Grady Trauma Project, individuals with a Grady delivery, 2011-2023, n = 704.

No type of trauma was associated with elevated risk of hypertension during pregnancy (Table 3) in either unadjusted models or after adjusting for parity, age, and education. Similarly, trauma recency was not associated with elevated risk of hypertension (Table 4). These results were maintained following incorporation of inverse probability of participation weights (Tables S2 and S3) and across specific diagnosis category (Table S4).

**Table 3.** Estimated associations between trauma events ever and by timing of exposure and hypertensive disorders of pregnancy,* Grady Trauma Project, individuals with a Grady delivery, 2011-2023, n = 700.

|  | HDP, unadjusted<br>Risk Ratio<br>(95% CI) | HDP, adjusted †<br>Risk Ratio<br>(95% CI) |
| --- | --- | --- |
| **Any trauma** |  |  |
| Any trauma, 0-9 | 1.05 (95% CI:0.86-1.23) | 1.01 (95% CI:0.83-1.19) |
| Any trauma, 10-19 | 0.79 (95% CI:0.63-0.96) | 0.8 (95% CI:0.64-0.96) |
| Any trauma, 20+ | 1.2 (95% CI:0.96-1.44) | 1.15 (95% CI:0.91-1.38) |
| **Non-interpersonal** | 1.14 (95% CI:0.91-1.38) | 1.12 (95% CI:0.89-1.34) |
| 0-9 | 1.18 (95% CI:0.94-1.42) | 1.17 (95% CI:0.9-1.44) |
| 10-19 | 0.87 (95% CI:0.71-1.03) | 0.87 (95% CI:0.71-1.03) |
| 20+ | 1.08 (95% CI:0.87-1.3) | 1.04 (95% CI:0.83-1.24) |
| **Violence** | 1.04 (95% CI:0.83-1.24) | 1.04 (95% CI:0.84-1.23) |
| 0-9 | 0.92 (95% CI:0.71-1.14) | 0.87 (95% CI:0.67-1.07) |
| 10-19 | 1.05 (95% CI:0.86-1.24) | 1.05 (95% CI:0.86-1.24) |
| 20+ | 1.05 (95% CI:0.83-1.26) | 0.99 (95% CI:0.79-1.19) |
| **Witnessing Violence** | 0.99 (95% CI:0.79-1.19) | 0.98 (95% CI:0.77-1.2) |
| 0-9 | 0.96 (95% CI:0.78-1.15) | 0.97 (95% CI:0.78-1.15) |
| 10-19 | 0.89 (95% CI:0.73-1.04) | 0.9 (95% CI:0.74-1.06) |
| 20+ | 1.03 (95% CI:0.82-1.25) | 1 (95% CI:0.79-1.21) |
| **Sexual trauma** | 1 (95% CI:0.79-1.21) | 0.97 (95% CI:0.78-1.15) |
| 0-9 | 0.99 (95% CI:0.75-1.24) | 0.94 (95% CI:0.72-1.17) |
| 10-19 | 1.01 (95% CI:0.79-1.22) | 1.03 (95% CI:0.82-1.24) |
| 20+ | 1.21 (95% CI:0.75-1.66) | 1.02 (95% CI:0.62-1.42) |

\*Hypertension inclusive of chronic hypertension, gestational hypertension, preeclampsia, or eclampsia
†Adjusted for parity (primiparous or multiparous), education (binary), age at delivery ( < 20 years, 20-34, 35+)

**Table 4.** Estimated associations between recency of exposure to trauma and hypertensive disorders of pregnancy,* Grady Trauma Project, individuals with a Grady delivery, 2011-2023, n = 700.

|  | HDP, unadjusted<br>Risk Ratio<br>(95% CI) | HDP, adjusted<br>Risk Ratio<sup>†</sup><br>(95% CI) |
| --- | --- | --- |
| **Non-interpersonal (n = 440)** |  |  |
| < 3 years prior to delivery | ref | ref |
| 3-10 years prior to delivery | 1.07 (95% CI:0.78-1.37) | 1.09 (95% CI:0.79-1.39<sup>‡</sup>) |
| 10 + years prior to delivery | 1.27 (95% CI:0.94-1.61) | 1.23 (95% CI:0.91-1.55) |
| **Violence (n = 398)** |  |  |
| < 3 years prior to delivery | ref | ref |
| 3-10 years prior to delivery | 1.09 (95% CI:0.75-1.44) | 1.08 (95% CI:0.74-1.42) |
| 10 + years prior to delivery | 1.43 (95% CI:1-1.86) | 1.36 (95% CI:0.93-1.79) |
| **Witnessing Violence (n = 525)** |  |  |
| < 3 years prior to delivery | ref | ref |
| 3-10 years prior to delivery | 0.81 (95% CI:0.56-1.06) | 0.82 (95% CI:0.56-1.07) |
| 10 + years prior to delivery | 1.2 (95% CI:0.89-1.51) | 1.15 (95% CI:0.83-1.46) |
| **Sexual trauma (n = 257)** |  |  |
| < 3 years prior to delivery | ref | ref |
| 3-10 years prior to delivery | 0.64 (95% CI:0.24-1.05) | 0.67 (95% CI:0.26-1.08) |
| 10 + years prior to delivery | 0.94 (95% CI:0.44-1.44) | 0.9 (95% CI:0.43-1.38) |

\*Hypertension inclusive of chronic hypertension, gestational hypertension, preeclampsia, or eclampsia
<sup>†</sup>Adjusted for parity (primiparous or multiparous), education (binary), age at delivery ( < 20 years, 20-34, 35+)
<sup>‡</sup> Adjusted for age and education only

## Discussion

In the current study, the type and timing of trauma exposure did not predict hypertension during pregnancy. This result was in contrast to our hypothesis and prior literature, yet robust to sensitivity analyses. This community sample experienced extremely high rates of trauma with the majority of surveyed individuals reporting exposure to direct experiences of violence and witnessing violence.^24^ For example, compared to an analysis of the 2011-2014 Behavioral Risk Factor Surveillance System data, participants reported nearly twice the burden of intimate partner violence (31.7% compared to 18.2%) and nearly twice the rate of sexual abuse (26.7% compared to 16.3%).^31^ Further, the burden of hypertension was nearly four times the national average of 15.9%(though similar to the burden experienced among the hospital as a whole).^32^ These null results suggest nuance to the growing body of work showing an association between trauma and adverse pregnancy outcomes.^16,33,34^

There are several possible explanations for the observed null results. First, it may be that the “unexposed” group was an unusual and otherwise high risk group. Supporting that, the “no trauma” group had a higher rate of chronic and gestational diabetes than the trauma exposed group, in contrast to prior literature. Second, it is likely that this population continued to experience notable levels of stress and trauma during their pregnancy that our survey was unable to capture, as supported by prior research in this population.^21^ This may have acted as a modifier of the association between lifetime trauma and hypertension during pregnancy. Third, it is possible that some threshold exists at which additional adversity does not confer additional cardiometabolic risk during pregnancy. Many of the prior studies of trauma and pregnancy outcomes were conducted in population-based samples with lower levels of ongoing social risks compared to this population.^34,35^ Possibly, ongoing social risks outweighed historic trauma in this population. There are likely many possible pathways (or sufficient component causes) by which someone may develop hypertension during pregnancy.^36^ It may be that individuals susceptible in this population have already completed the sufficient component causes regardless of trauma exposure through other physical and social risks. For example, risks from pollution exposure, low economic resources, poor metabolic health, family history of hypertension, and lack of primary care prior to pregnancy are common in this sample, any combination of which may have resulted in HDP. Fourth, primiparous and younger women were underrepresented in this sample. It may be that the impacts of life course trauma on pregnancy outcomes is mediated by parity and age at first pregnancy.^37,38^ We are unable to tease that out given that we only include a single delivery per participant and not a full reproductive history. This is not to state that trauma had no impact on the lives of participants. Notably, we observed a strong positive relationship between trauma burden and psychiatric symptoms, consistent with prior literature.^39,40^

The results of the current study should be interpreted in light of its limitations. First, the study population represents a selected sample with potential overrepresentation of individuals with prior mental health morbidity. However, inclusion of inverse probability of selection weights did not change associations meaningfully. Second, we were unable to fully control for childhood socioeconomic status (SES) and used a proxy and potential mediator, high school graduation. This may bias results towards the null. However, results were similar in adjusted and unadjusted models, suggesting that this was unlikely to be a major source of bias. Alternately, this proxy may have imperfectly captured SES, resulting in residual confounding. However, this study sample represents a relatively homogenous, low-SES sample, so we do not believe there would have been sufficient variation in childhood SES to substantially bias the results. Third, trauma was self-reported and recall of trauma is known to change over the life course, including during pregnancy.^41^

Exposure to traumatic events tends to cluster along the lines of socioeconomic position. Individuals who experience poverty, structural and interpersonal discrimination, and are of minoritized racial, national, or ethnic backgrounds are more likely to experience trauma across the life course.^5–8,42^ The Weathering Hypothesis and other theoretical frameworks suggest that the disproportionate burden of stress and trauma among individuals with minoritized identities may drive racial disparities in perinatal outcomes.^18^ Our current results demonstrate the challenge of understanding this relationship using data collected only at the time of pregnancy and within single hospitals. At the time of their pregnancies, participants in our study had experienced more reported trauma than most people will ever experience. Many reported experiencing trauma beginning at very early ages. Due to a highly segmented healthcare market for obstetric care, people with Medicaid and without insurance may have a smaller range of choice in delivery hospitals, concentrating individuals with physical and social challenges in a single care site.^43,44^ To better understand how weathering occurs, we need longitudinal data. To prevent the toxic consequences of weathering on maternal and infant health, prevention of trauma and early interventions are critical. These results also highlight the need to incorporate socioeconomic diversity into studies of trauma and health outcomes. While trauma was not a predictor of hypertension in this pregnant population, the population experienced an extraordinarily high burden of trauma, hypertension, post-traumatic stress symptoms, and depressive symptoms. Universal perinatal screening and referral for mood disorders and PTSD and trauma-informed obstetric care may be beneficial for maternal and infant health.^45–47^ Future research and clinical interventions focused on improving maternal care for communities experiencing high burdens of trauma are critical to improving maternal health outcomes.

## Data Availability

Deidentified data from the two individual datasets linked in this article is available upon request to the specific studies (Grady Trauma Project and the Grady Obstetric and Gynecologic Outcomes database). Linked identified data is not available.

